# Viral shedding and symptom severity across populations during acute COVID in the ACTIV-2 study

**DOI:** 10.64898/2026.01.31.26345293

**Authors:** Evelyn Kung, Rinki Deo, Manish C. Choudhary, Kara W. Chew, Teresa H. Evering, Rachel Bender Ignacio, Prasanna Jagannathan, James P. Flynn, James Regan, Carlee Moser, Mark J. Giganti, Michael D. Hughes, Justin Ritz, Arzhang Cyrus Javan, Alexander L. Greninger, Upinder Singh, William Fischer, Eric S. Daar, David A. Wohl, Joseph J. Eron, Judith S. Currier, Robert W. Coombs, Davey M. Smith, Jonathan Z. Li

## Abstract

To evaluate the impact of sex on acute SARS-CoV-2 infection, 668 participants from the ACTIV-2/A5401 study were followed over a 28-day period. A primary analysis was performed on the 469 participants who had quantifiable viral loads at baseline. Male and female participants had comparable nasal SARS-CoV-2 RNA levels at study entry and throughout follow-up. However, sex-specific differences in viral shedding emerged when stratified by duration of symptoms. In the first three days from symptom onset, female participants exhibited higher nasal SARS-CoV-2 RNA levels than males, but lower viral RNA levels thereafter. The higher viral RNA levels in females during the earliest phase of acute COVID-19 was seen even after adjusting for age, race and region of enrollment. Female participants also tended to have higher symptom scores across days since symptom onset but no significant correlation was observed between nasal SARS-CoV-2 RNA levels and symptom score regardless of sex. These findings highlight the impact of sex on both viral shedding and symptom dynamics and underscore the importance of considering time since symptom onset when evaluating respiratory virus antiviral therapies in clinical trials.

## Introduction

The COVID-19 pandemic, caused by Severe Acute Respiratory Syndrome Coronavirus 2 (SARS-CoV-2), has had an unprecedented impact on global mortality [1]. Previous research has established the relationship between older age and the presence of comorbidities (e.g., diabetes, hypertension) with increased COVID-19 morbidity and severity [2-4]. There is a need to further understand the role of sex as a modifier of viral shedding and symptoms during acute COVID-19 [5, 6]. Male individuals have been well-described to be at greater risk of severe COVID-19. One analysis reported that ICU admission rates in male patients were twice as high as in females, identifying male sex as an independent risk factor for mortality [7]. Similarly, a study of hospitalized patients in the United Kingdom showed that males were placed in more severe symptom categories compared to females [8]. How SARS-CoV-2 RNA decay and symptom severity during acute mild-moderate COVID-19 might differ by sex would benefit from additional exploration. Prior work in the ACTIV-2/A5401 study reported some intriguing findings, including that there may be some differences in SARS-CoV-2 RNA levels at baseline by race and that viral decay may differ by sex [9]. In this study, we expanded upon this analysis including additional placebo-recipients of ACTIV-2/A5401, performing a concurrent analysis of how symptom scores differ by sex and evaluating how our findings differed when analyzed by study day (as most SARS-CoV-2 treatment trials do) and also by days since symptom onset.

## Methods

ACTIV-2/ ACTG (Advancing Clinical Therapeutics Globally) A5401 is a randomized, placebo-controlled platform trial designed to evaluate the safety and efficacy of investigational agents for the treatment of non-hospitalized adults with mild-to-moderate COVID-19 [9, 10]. The analysis included data from 668 participants enrolled between August 27th, 2020, and August 31st, 2021, who received placebo in the ACTIV-2/A5401 study and recruited from treatment centers across seven countries (United States, Brazil, South Africa, Mexico and Argentina). Participants were symptomatic non-hospitalized adults ≥18 years of age. We performed a primary analysis for 469 participants with quantifiable baseline SARS-CoV-2 viral loads and a secondary analysis that included participants who had undetectable viral loads at study entry.

All participants were enrolled in the study within 11 days since symptom onset (DSSO). The protocol was approved by a central institutional review board, Advarra (Pro00045266) and participants provided written informed consent before undergoing study procedures. All participants self-collected anterior nasal (AN) swabs at days 0, 3, 7, 14, and 28 for quantitative SARS-CoV-2 RNA testing as previously described [9, 11]. Viral decay during the early phase of infection was estimated using longitudinal viral load measurements from study entry up to, but not including, the first visit at which viral load fell to ≤ LLOQ (1.7 log□□). Participants with fewer than two viral load measurements within this interval were excluded from slope estimation. For each participant, viral decay was quantified by fitting a linear regression of log□□ (viral load) versus study day, with the slope representing the rate of viral load decline (log□□ units per day).

Total symptom scores were calculated based on a 29-day diary completed by the participants for 13 targeted symptoms [10]. The targeted symptoms were feeling feverish, cough, shortness of breath or difficulty breathing, sore throat, body pain or muscle pain or aches, fatigue, headache, chills, nasal obstruction or congestion, nasal discharge, nausea, vomiting, and diarrhea. Each symptom was recorded daily by the participant as absent (score 0), mild (1), moderate (2), or severe (3). Total symptom score was calculated for a given day by summing the scores for the 13 symptoms (possible range of 0-39).

Categorical variables were summarized using frequencies and percentages, and between-group differences were evaluated using either chi-squared test or Fisher’s exact test as appropriate. Continuous variables were summarized with median and interquartile ranges and compared with non-parametric methods (Wilcoxon rank-sum test to compare two groups. R (4.3.0) was used for statistical analyses. Two-tailed tests were used for all the analyses and P<0.05 was considered statistically significant.

## Results

A total of 469 ACTIV-2/A5401 trial participants who received placebo were studied, including 220 males (47%), 249 females (53%), 210White (45%), 41 Black (9%) and 183 Hispanic (39%). The median (Q1, Q3) age for males was 48 years (38.75, 57) with females having a similar distribution of 48 (36, 55). Male and female participants had similar days since symptom onset at the time of enrollment (male vs female, median [Q1, Q3]: 5.5 [3,7] vs 6 [4,7]). Further participant demographics can be found in Table 1.

**Table 1.** Demographic characteristics of participants stratified by sex. Statistical analysis was performed using Wilcoxon rank-sum tests for quantitative variables and Chi squared tests for categorical variables. **Abbreviations:** SS, Symptom Score; VL, viral load; ns, non-significant

| Characteristic | Male<br>(n=220 ) | Female<br>(n=249 ) | P value |
| --- | --- | --- | --- |
| **Median age (Q1, Q3)** | 48 (38.75,57) | 48 (36,55) | 0.24 |
| **Country, n (%)** |  |  |  |
| ARG | 13 (5.6) | 19 (7.6) | 0.93 |
| BRA | 8 (3.6) | 8 (3.2) |  |
| MEX | 1 (0.45) | 1 (0.40) |  |
| USA | 181 (82.3) | 199 (79.9) |  |
| ZAF | 17 (7.7) | 22 (8.8) |  |
| **Symptom score at study entry (Q1, Q3)** | 9 (6,13) | 11 (7,16) | <0.01 |
| **Median AN SARS-CoV-2 viral load at study entry in $\log_{10}$ copies/mL (Q1,Q3)** | 4.96 (3.87, 6.48) | 5.18(3.74,6.35) | 0.91 |
| **Median days from symptom onset to study entry (Q1, Q3)** | 5.5 (3,7) | 6 (4,7) | 0.36 |

We first performed an analysis of SARS-CoV-2 viral load by study day for participants with detectable VL at baseline. There were no significant differences in the proportion of male versus female participants with Vl<LLoQ at the time of study entry (29% F vs. 31%M, p=0.43). While male and female participants exhibited similar baseline levels of nasal SARS-CoV-2 RNA levels at study entry, females had a significantly higher viral load at study day 3 (P<0.01; Figure 1A). There were no significant differences in viral load by sex at study days 7, 14, or 28.

**Figure 1.**
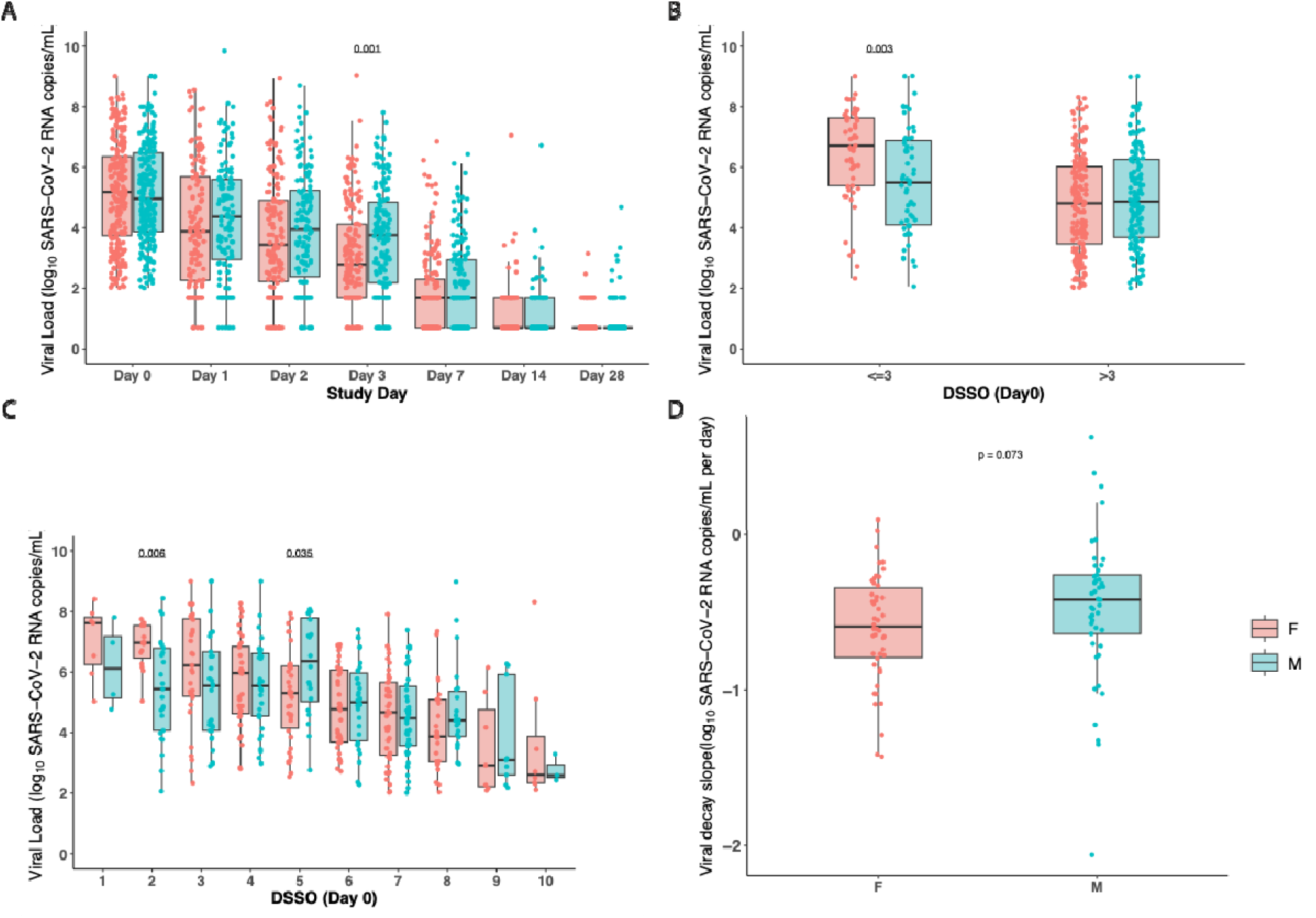
Nasal SARS-CoV-2 viral load (VL) comparisons between males and females. (A) VL by study day. (B) VL at enrollment (Day 0) stratified by ≤3 vs >3 days since symptom onset (DSSO). (C) VL at enrollment (Day 0) further categorized by individual DSSO days (1– 10). (D) Viral decay at DSSO Day 0. Boxplots represent the interquartile range (IQR; 25th–75th percentile), with the horizontal line indicating the median. Individual data points are overlaid as dots. *P*-values were calculated using Wilcoxon rank-sum tests.

However, analysis by study day does not take into account the timing of symptom onset for each participant. When nasal RNA levels at the time of study entry was analyzed by accounting for the days since symptom onset (DSSO), we did note that early in the disease course (≤3 days from the time of symptom onset), there was a difference in the nasal viral RNA levels by sex: female participants had significantly higher viral loads than male participants at study entry (female vs male: median 6.7 vs. 5.5 SARS-CoV-2 RNA log_10_ copies/mL, P<0.01; Figure 1B). Later from symptom onset (DSSO≥5 days), levels of SARS-CoV-2 was generally similar or lower in female compared to male participants (Figure 1C). In a multivariable analysis of participants who entered the study ≤3 days from the time of symptom onset, female participants had higher viral RNA levels (P=0.004) after adjusting for age, race, and geographic region of enrollment (Supplemental Table 1). We also performed a secondary analysis in which we analyzed all participants (668 participants), including those with undetectable SARS-CoV-2 RNA levels at the time of study entry. Similar trends in higher viral loads in female participants earlier after study entry (≤3 DSSO) was also observed (Supplemental Figure 1). To further evaluate whether these differences translated into differences in viral clearance, we next assessed early viral decay rates. Viral decay showed a trend toward faster decline in female compared with male participants, although this difference did not reach statistical significance (median viral decay for F vs M: -0.59 vs -0.42 log_10_/day, P=0.07; Figure 1D).

Similar to the viral load analysis above, we also performed an analysis of symptom scores between female and male participants at the time of study entry by days since study onset. In general, female participants had higher levels of symptom scores across DSSO, with significantly higher levels found only at day 8 after symptom onset (Figure 2). The results were similar in the secondary analysis that included participants with undetectable nasal viral RNA at study entry (Supplemental Figure 2). We also analyzed the relationship between viral load and symptom score and found no significant correlation between SARS-CoV-2 RNA levels and symptom severity by sex (Figure 3) at the time of study entry.

**Figure 2.**
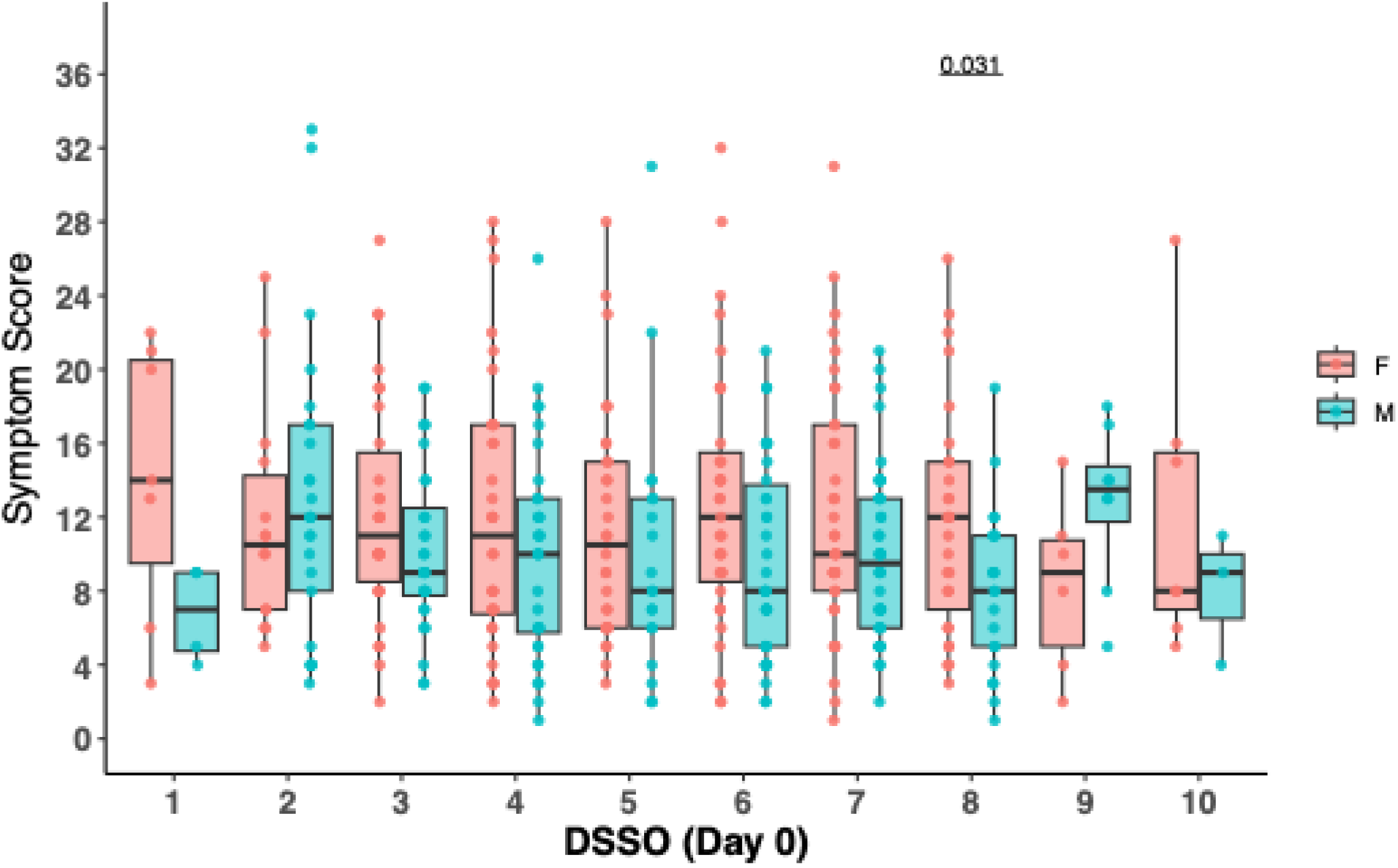
Comparison of male and female symptom scores by days since symptom onset (DSSO). Data are shown as boxplots representing the interquartile range (IQR; 25th to 75th percentile) with the median as a horizontal line within the box. Individual data points are shown as dots. P-values were calculated using Wilcoxon rank-sum tests.

**Figure 1.**
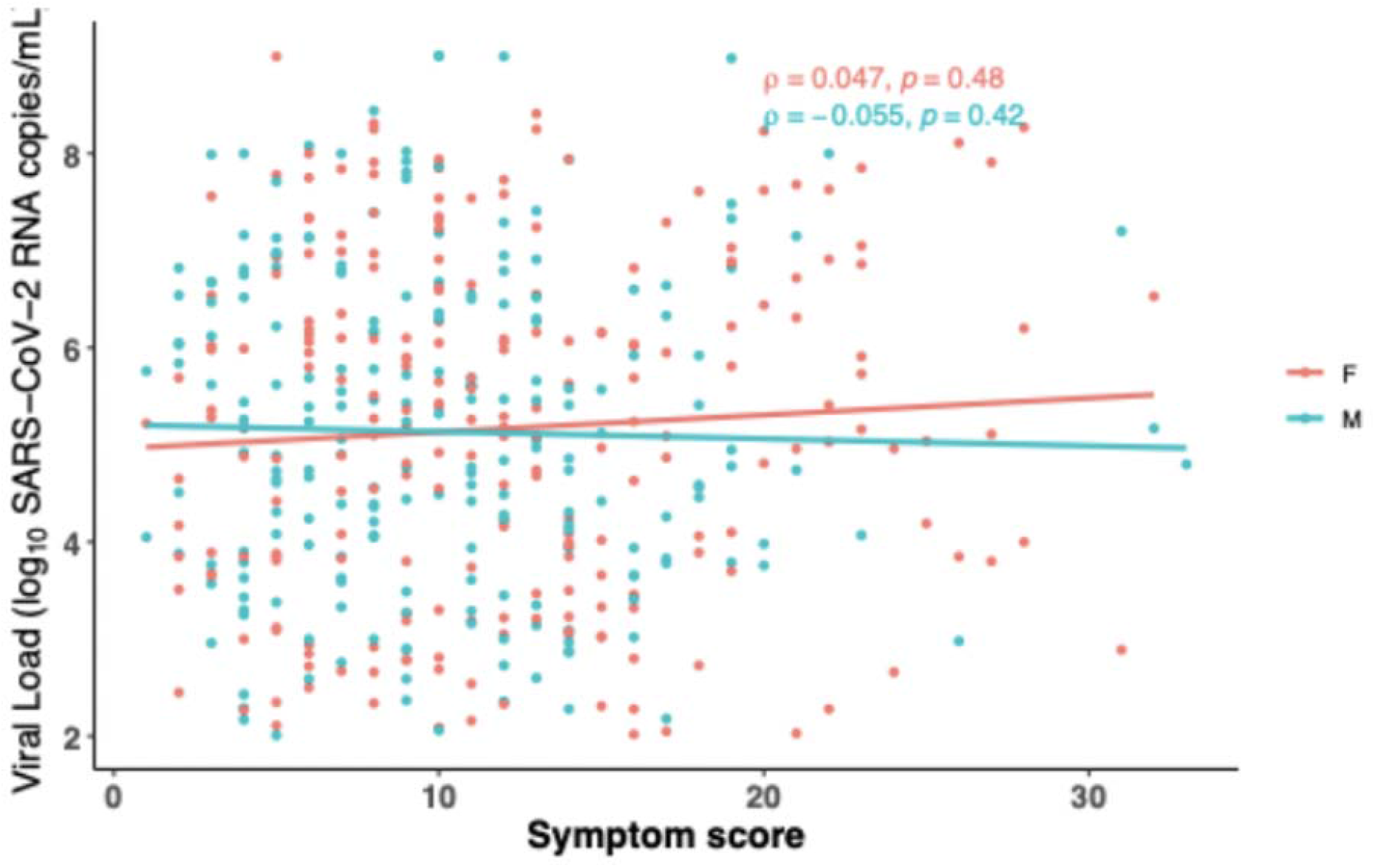
Comparison of Symptom Score (SS) and nasal SARS-CoV-2 viral load (VL) stratified by sex. Each dot represents an individual participant. Simple linear regression was used to generate lines of best fit, and Spearman’s rank correlation was performed to assess statistical significance. P-values less than 0.05 were considered significant.

## Discussion

This study examined the relationship how sex may affect levels of nasal SARS-CoV-2 RNA and symptom severity. Using data from a rigorously performed clinical trial (ACTIV-2) that included a quantitative SARS-CoV-2 viral load assay, we found that sex-specific differences in viral shedding were more evident when considering the duration of symptoms than just by study day. In the first three days from symptom onset, female participants had higher nasal SARS-CoV-2 viral loads compared to men. We also found that women generally had higher symptom scores at study entry regardless of DSSO but that there were no significant correlation between nasal SARS-CoV-2 RNA levels and symptom score regardless of sex. A secondary analysis of all participants, including those with undetectable SARS-CoV-2 RNA levels at study entry, demonstrated concordant results.

Within the field of virology, there have been numerous examples of how biological sex may impact viral infections. In HIV, female sex has been found to impact the extent of plasma viremia, which is likely modulated by the estrogen hormone and its effects on T cell activation and HIV reservoir activity [12, 13]. In the influenza field, human challenge experiments have shown that women appear to have a greater number of symptoms than men, although the number of days of viral shedding did not differ by sex [14]. Studies of COVID-19 have reported disparate results, including some studies reporting sex-based differences in viral shedding [15, 16], while others have not [17]. However, strengths of this study include the rigorous clinical trials structure and the use of quantitative SARS-CoV-2 viral load assay, which is not readily available outside of a clinical trials setting. A prior study of ACTIV-2 clinical trials participants had reported no significant difference in nasal SARS-CoV-2 RNA levels at study entry between female and male participants, although there appeared to be a greater decline in viral RNA in female participants [9]. Our results show the importance of taking into account the timing of symptom onset when analyzing viral shedding and symptoms, given the wide range of symptom duration of trial participants at the time of study entry.

In addition to its role in the female reproductive cycle, estrogen is also a key regulator of inflammation within the respiratory tract and modulates both the innate and adaptive immune responses [18, 19]. Intriguingly, there is also evidence that the SARS-CoV-2 spike protein binds the estrogen receptor, inducing estrogen-receptor-dependent biological effects and potentiating estrogen effects in alveolar macrophages and other cell types [20]. Prior studies have reported discordant findings on the relationship between concurrent SARS-CoV-2 RNA shedding and symptom scores [21, 22], although a human challenge experiment found no correlation between viral load and symptoms [23]. We have now extended those findings to show a lack of a significant correlation between viral RNA levels and symptom scores in both female and male participants.

Limitations to this study include that these participants were largely enrolled during the early phases of the COVID-19 pandemic and it is unclear how changes in the omicron variants and host immunity may affect these results. In addition, it is unclear why a subset of individuals had SARS-CoV-2 RNA levels below the limit of quantification at the baseline timepoint. It is possible that a subset of participants may have had rapid viral clearance prior to study enrollment, although we found no significant differences in the proportion of females and men who had undetectable viral RNA at baseline and a secondary analysis including all participants demonstrated concordant results.

In summary, our analysis of a rigorously performed clinical trial highlight sex-based variations in nasal viral RNA shedding and symptom scores. These results emphasize the importance of considering time since symptom onset when analyzing clinical trials of antiviral treatments and suggest the need for further research to better understand how estrogen and sex influence the progression and outcomes of respiratory tract infections.

## Data Availability

All data produced in the present work are contained in the manuscript

## Acknowledgements

The authors thank the study participants, site staff, site investigators, and the entire ACTIV-2/A5401 study team.

## Financial support

This work was supported by the National Institute of Allergy and Infectious Diseases of the National Institutes of Health under Award Number UM1AI068636, UM1AI068634, and UM1AI106701. The content is solely the responsibility of the authors and does not necessarily represent the official views of the National Institutes of Health.

## Conflicts of Interests

Authors declare no conflict of interest.

---

**Supplemental Table 1.** Multivariate analysis of SARS-CoV-2 viral load at study entry in those with quantifiable results and <=3 days from symptom onset.

| Variable | $\beta$ | 95% CI | p-value |
| --- | --- | --- | --- |
| **AGE** | 0.00 | -0.02, 0.03 | >0.9 |
| **SEX - F** | — | — | — |
| **SEX - M** | -0.92 | -1.5, -0.30 | 0.004 |
| **Race - BLACK** | — | — | — |
| **Race - HISPANIC** | -0.60 | -2.2, 1.0 | 0.5 |
| **Race - OTHER** | 1.2 | -0.55, 2.9 | 0.2 |
| **Race - WHITE** | -0.06 | -1.6, 1.7 | >0.9 |
| **Country/Region - North America** | — | — | — |
| **Country/Region - South Africa** | -0.57 | -2.3, 1.2 | 0.5 |
| **Country/Region - South America** | 0.35 | -0.96, 1.7 | 0.6 |
| Abbreviations: CI = Confidence Interval, $\beta$ coefficient with 95% Confidence Interval | | | |

**Supplementary Figure 1.**
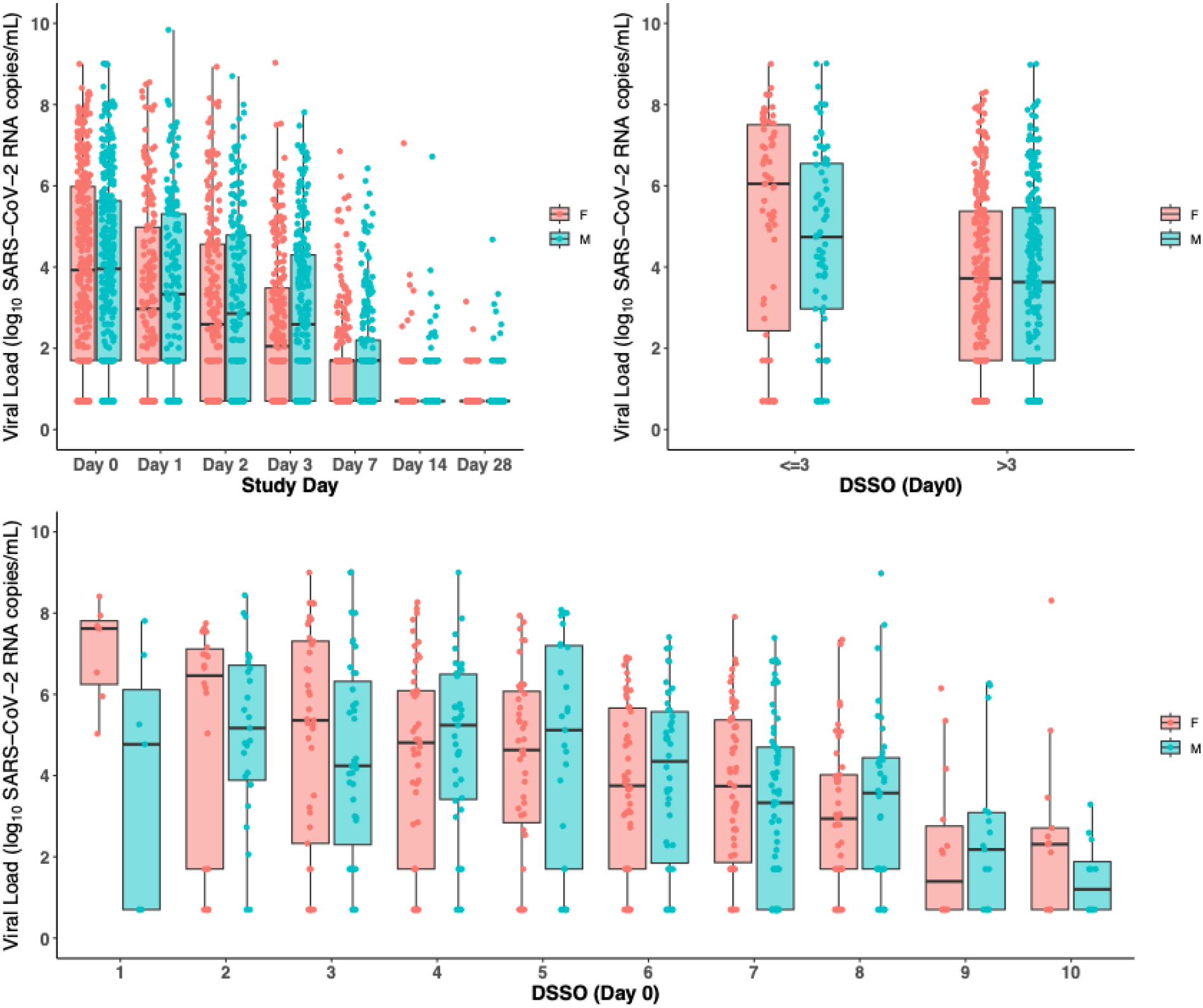
Nasal SARS-CoV-2 viral load (VL) comparisons between males and females for 668 participants, including those with undetectable viral RNA levels at time of study entry. (A) VL by study day. (B) VL at enrollment (Day 0) stratified by ≤3 vs >3 days since symptom onset (DSSO). (C) VL at enrollment (Day 0) further categorized by individual DSSO days (1–10). Boxplots represent the interquartile range (IQR; 25th–75th percentile), with the horizontal line indicating the median. Individual data points are overlaid as dots. *P*-values were calculated using Wilcoxon rank-sum tests.

**Supplemental Figure 2.**
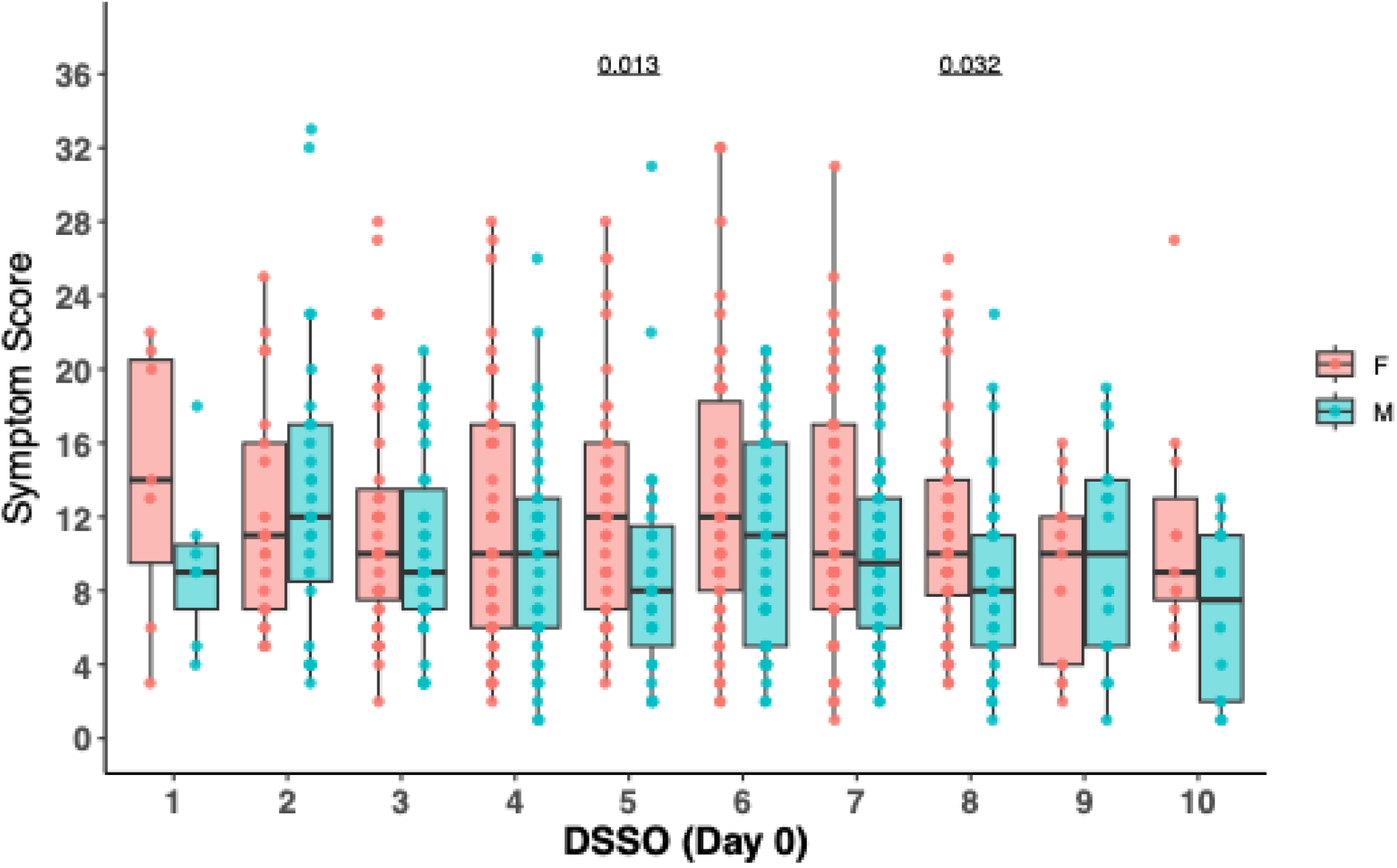
Comparison of male and female symptom scores by days since symptom onset (DSSO) from secondary analysis including all participants. Data are shown as boxplots representing the interquartile range (IQR; 25th to 75th percentile) with the median as a horizontal line within the box. Individual data points are shown as dots. P-values were calculated using Wilcoxon rank-sum tests.

## Notes

### Competing Interest Statement

The authors have declared no competing interest.

### Clinical Trial

NCT04518410

### Author Declarations

The protocol was approved by a central institutional review board, Advarra (Pro00045266) and participants provided written informed consent before undergoing study procedures.

